# Common genetic alterations of *SPOP-MATH* domain in prostate cancer tissues and association with pathological tumor characteristics^*^

**DOI:** 10.1101/2021.09.09.21263201

**Authors:** Berjas Abumsimir, Mohammed Mrabti, Abdelilah Laraqui, Imane Saif, Maryame Lamsisi, Youssef Ennaji, Ahmed Ameur, Saad Ibnsouda Koraishi, Moulay Mustapha Ennaji

## Abstract

*SPOP* gene has a critical role in prostate cancer development and found high mutated in the prostate tumor through various populations. MATH domain represents an important site for *SPOP*-DNA linkage and other sensitive gene-gene interactions. To investigate the genetic alterations of the MATH domain of *SPOP* gene in prostate cancer biopsies and correlation with clinical and pathological parameters; DNA samples from 50 prostate cancer tissues were genotyped and confirmed by Sanger sequencing. The frequency and distribution of high frequent mutations were determined and correlated with the patient’s tumor characteristics. Among 50 samples 34 (68%) were carrying one or more common mutations. Novel frame shift deletion mutation: c.255delA (p.Leu86Phefs) was detected in eight patients (16%), in addition to five novel missense mutations with moderate frequency (6%) namely: c.209G>C (p.Arg70Pro), c.215A>C (p.Asn72Thr), c.334G>A (p.Glu112Lys), c.373T>C (p.Phe125Leu), and c.388G>A (p.Asp130Asn), All missense mutations located in MATH domain. The effects of novel mutations described in the MATH domain are uncertain. No significant differences between carriers and noncarriers of common mutations detected regarding Gleason score, prostate-specific antigen concentration PSA, and tumor stage [p > 0.05]. Clinical significance of mutations detected on prostate tumors progression can be investigated in future analysis. Our findings revealed novel *SPOP* alterations in prostate cancer tissues probably associated with cancer development.

## Introduction

Prostate tumors are generally among the most common types of tumors among men in the world, according to the statistics issued by the competent authorities the mortality rate counting 1,276,106 new cases and causing 358,989 deaths in 2018 worldwide [1]. Certain gene variants contribute to an elevated risk of prostate cancer. Various alleles appear repeatedly mutant in specific genes in prostate patients. Speckle-type POZ (*SPOP*) gene encodes a protein that modulates the transcriptional repression functions of death-associated protein 6 (*DAXX*). Several Mutations in *SPOP* lead to a type of hereditary prostate tumor which involved 10-14.4% of all prostate tumors across different demographic and ethnic origins. There is no correlation between SPOP mutations with ethnicity, clinical, or pathologic parameters. The expression and mutational status, defining a new biotype of Prostate cancer associated with a worse prognosis as mentioned elsewhere [2–6]. Recently *SPOP* mutations suggested to drives prostate tumorgenesis in part through genomic instability; *SPOP* somatic mutations also linked with ETS rearrangements and TMPRSS2-ERG fusions in prostate cancer tissues [7–10]. Moroccan population is a unique race mixed North African populations. In this population, Prostate cancer considers the second most common cancer in men with 3990 new cases in 2018 [11]. This study aimed to investigate the prevalence of the common mutations of conserved domain MATH of *SPOP* gene that contributed to prostate tumor clinical characteristics.

## Materials and methods

### Prostate cancer biopsies

Fifty fresh prostate biopsies were derived from fifty men who were evaluated at Mohammed v teaching hospital of Rabat city between June 2017, and February 2019. Ethical standards have been respected before and during biopsies preparation, including. Required Ethical approval was obtained from the committee of biomedical research ethics in Morocco (No.: 3/2018/30 April/2018). Histological evaluation has been already obtained to diagnose them of the presence of prostatic adenocarcinoma. Samples were stored after obtaining the biopsies according to clinical protocols by physicians directly. Fifty specimens were directly extracted. The following clinico-pathological parameters were recorded: age at diagnosis, date of biopsy, Gleason score, PSA concentration, and radical prostatectomy treatment. DNA processing and genotyping were took place in oncology and virology laboratory in the faculty of sciences and techniques at Mohammedia, Morocco.

### DNA processing and Genotyping

Approximately 25 mg of every prostate biopsy was used as source of genomic DNA, according the instruction of ready kit: pure link Invitrogen genomic DNA mini kit®, Thermo Fisher USA. DNA was quantified using the Nanodrop spectrophotometer. Samples with a DNA concentration of 20-50 ng/μl or above were selected to perform the polymerase chain reaction. The extracted DNA was quantified Nanodrop spectrophotometer. The concentration of DNA of 20-50 ng/μl or more were selected to be a template of DNA amplification by polymerase chain reaction. Exons six and seven of the *SPOP* gene were targeted by amplification using specific primers described elsewhere [2], in a 25 μl PCR reaction containing genomic DNA (8ng), 2× Taq PCR master mix kit Qiagen USA,1 μl of 10 μmol specific sense and antisense primers. Perkin Elmer 2400 Thermal Cycler®, CA, USA, was used to perform PCR amplification. The amplification programs for exon six and seven were as follows: initial denaturation at 94°C for 3 minutes, followed by 35 cycles of: denaturation at 94°C for one minute, annealing at 52°C for exon 6 and 48°C for exon 7, for one minute, extension at 72°C for one minute and final extension at 72°C for 10 minutes. In every PCR process a ultra pure water was used as negative control. Amplicons sizes of 170 for exon six and 240 bp for exon seven were confirmed by gel electrophoresis for 1.5 hours at 70 volts on 2% agarose. Preparing the sequencing process, PCR products were purified using Express PCR Product Cleanup ExoSAP-IT™ kit. Bidirectional Sanger sequencing reaction was performed using suitable ready kit: BigDye Terminator v1.1 Cycle Sequencing, using the same primers used in PCR by Genetic analyzer 3130 Applied Biosystems, and Last purification by BigDye XTerminator® Purification Kit, according to instructions of the manufacture. Sequencing processes were performed in innovation centre at Sidi Mohammed bin Abdullah University of fez, Morocco.

### Mutations identification and registration

Sequencing results processing and mutations detection were performed using MEGA software and Nagahama mutation finder server [12]. The resulted mutations were compared with *Refseq* reference: (NM_001007226.1). Common missense, nonsense, frameshift mutations effects were discussed in results, full *SPOP* molecular alterations results can be requested from the authors directly. The novel variants detected in this study were submitted to the U.S. National Library of Medicine (NLM), ClinVar database and successfully accredited as clinical relevance variants, Condition ID type: MedGen, Condition ID value: (C0376358: Malignant tumor of the prostate), under accession numbers: (SCV000998584**-**SCV000998743). SPOP DNA sequences resulted were submitted successfully to GenBank library under accessions: (MN606081-MN606100). The variants have been verified to be compatible with HGVS nomenclature standards by https://mutalyzer.nl [13].

### Statistical analysis

The samples were classified according to common mutations detected to into two groups: carriers of common mutations and noncarriers. The clinco-pathological parameters for two groups were recorded. The following tumor parameters were used for statidtical comparisions: Gleason score, prostate specific antigen (PSA) concentrations, Tumor stage, and age at diagnosis, and radical prostatectomy treatment. The statistical correlations were based on 2 × 2 or more chi-squared analysis or Fisher’s exact test to test for statistical significance. The unknown data were excluded, and minor mutation frequency was determined based on the total number of mutations among all study subjects. Analyses were conducted using the Minitab 17 (Coventry, United Kingdom). The P values of 0.05 or less were considered statistically significant.

### *In silico* prediction of novel missense variant affects

The missense variants usually have an unknown effect on protein function; the computational prediction tools confer a prediction probability to explore the negative effects of missense variants on the targeted protein. SIFT Blink and Poly Phen 2, two of the most efficient tools for these purposes. SIFT is a sequence homology-based tool classifying the missense variants as tolerated (if the score is more than 0.05) or affect the protein functions [14]. While the Poly Phen2 calculates the PSIC (position-specific independent count) score and considers the missense variants as one of three categories: probably damaging the protein function, possibly damaging, or benign [15]. The native Speckle type BTB/POZ, accession: NP_001007226.1, 374 amino acids, and mutated SPOP protein’s sequences were submitted as fasta format along with the positions of the native and mutant substitutions amino acids to SIFT and Poly Phen2 servers.

## Results

The clinical characteristics of study subjects are shown in Table 1. Among 50 patients, thirty-seven (74%) were diagnosed with prostate cancer at an age of more than 60 years old, and thirty-three (66%) of them have a concentration of prostate-specific antigen PSA equal to 10 mg /ml or more. Twenty-five cases (50%) had a Gleason score of 7 or more, and only eight cases were at the pathological tumor stage of T3 or T4 at the time of biopsy. Sixteen (32%) of patients treated radical prostatectomy, in addition to 10 cases (20%) were alcoholics, and eighteen subjects (36%) were regular smokers.

**Table1:** Clinical characteristics of prostate cancer patients.

| Characteristic | n (%) |
| --- | --- |
| Total cases | 50 |
| Age at diagnosis/surgery |  |
| ≤60 years | 7 (14) |
| >60 years | 37 (74) |
| Unknown | 6 (12) |
| Preoperative PSA, ng/mL |  |
| <2.5 | 1 (2) |
| 2.5–10 | 10 (20) |
| ≥10.0 | 33 (66) |
| Unknown | 6(12) |
| Pathological Gleason score |  |
| <7 | 11 (22) |
| 7 (3 + 4) | 6 (12) |
| 7 (4 + 3) | 8 (16) |
| >7 | 11 (22) |
| Unknown | 14 (28) |
| Pathological T-stage |  |
| pT1 | 14 (28) |
| pT2 x | 17 (34) |
| pT3 x | 3 (6) |
| pT4 | 5 (10) |
| Unknown/pTx | 10 (20) |
| Radical prostatectomy |  |
| Yes | 16 (32) |
| No | 21 (42) |
| Unknown | 13 (26) |
| Alcohol consumption |  |
| Yes | 10 (20) |
| No | 32 (64) |
| Unknown | 8 (16) |
| Smoking |  |
| Yes | 18 (36) |
| No | 24 (48) |
| Unknown | 8 (16) |

The mutations were arranged firstly according to genotype, then by genome location, the effect on the amino acids, and frequency rate. Most frequent frameshift deletion mutations were: c.255delA (p.Leu86Phefs) in eight patients (16%), c.247delT (p.Tyr83Thrfs) in seven patients (14%) and c.354delG (p.Ser119Valfs) in six patients (12%). In addition to five missense mutations with same frequency (6%), namely: c.209G>C (p.Arg70Pro), c.215A>C (p.Asn72Thr), c.334G>A (p.Glu112Lys), c.373T>C (p.Phe125Leu), and c.388G>A (p.Asp130Asn). All these missense mutations mentioned are located in the MATH domain. Notably, one high frequent intronic mutation: c.480+38G>A, detected in eight patients (16%). Furthermore, synonymous substitution mutation: c.327C>T of Alanine at 109 loci with a high frequency of five prostate cancer patients (10%). The most common alteration revealed, genome locations, and phenotypes are given in Table 2.

**Table2:** Most frequent *SPOP* variants detected in prostate cancer biopsies. Reference genome database: (GRCh38), Human genome variation society (HGVS) Nomenclature. *Nucleotide and amino acid positions are based upon a coding sequence for Refseq accession NM_001007226.1.

| Genotype | CDS change* | Genome location | Protein change | Freq n=50 (%) |
| --- | --- | --- | --- | --- |
| Frameshift |  |  |  |  |
|  | c.247delT | Chr17:49619339 | p.Tyr83Thrfs | 7(14%) |
|  | c.255delA | Chr17:49619331 | p.Leu86Phefs | 8(16%) |
|  | c.269delT | Chr17:49619317 | p.Leu90Argfs | 3(6%) |
|  | c.274delA | Chr17:49619312 | p.Ser92Alafs | 4(8%) |
|  | c.276_277insC | Chr17:49619309 | p.Cys93Leufs | 3(6%) |
|  | c.284delA | Chr17:49619302 | p.Lys95Argfs | 3(6%) |
|  | c.315delC | Chr17:49619271 | p.Ile106Serfs | 3(6%) |
|  | c.354delG | Chr17:49619107 | p.Ser119Valfs | 6(12%) |
|  | c.360delA | Chr17:49619101 | p.Gln120Hisfs | 3(6%) |
| Missense |  |  |  |  |
|  | c.209G>C | Chr17:49619377 | p.Arg70Pro | 3(6%) |
|  | c.215A>C | Chr17:49619371 | p.Asn72Thr | 3(6%) |
|  | c.334G>A | Chr17:49619252 | p.Glu112Lys | 3(6%) |
|  | c.373T>C | Chr17:49619088 | p.Phe125Leu | 3(6%) |
|  | c.388G>A | Chr17:49619073 | p.Asp130Asn | 3(6%) |
| Intronic |  |  |  |  |
|  | c.353-11C>T | Chr17:49619119 | NA | 3(6%) |
|  | c.353-48C>A | Chr17:49619156 | NA | 3(6%) |
|  | c.480+38G>A | Chr17:49618943 | NA | 8(16%) |
| Synonymous |  |  |  |  |
|  | c.327C>T | Chr17:49619259 | p.Ala109= | 5(10%) |
|  | c.342C>G | Chr17:49619244 | p.Thr114= | 3(6%) |
|  | c.369T>C | Chr17:49619092 | p.Tyr123= | 2 (4%) |
|  | c.381A>G | Chr17:49619080 | p.Gln127= | 3(6%) |
|  | c.384C>G | Chr17:49619077 | p.Gly128= | 3(6%) |
|  | c.453T>G | Chr17:49619008 | p.Pro151= | 4(8%) |

The pathological and clinical parameters were compared between carriers and noncarriers of *SPOP* high frequent mutations in Table 3. High Gleason (more than 7) scores of prostate cancer among carriers were detected in three cases (less than 10%), while in noncarriers were five cases (31%) [P value =0.1944]. In contrast, nine (26%) cases among *SPOP* mutations carriers were less than 7, and eleven cases (32%) of the score were equal to 7. No significant differences between carriers and noncarriers regarding prostate-specific antigen. Numbers of cases of high PSA concentrations (PSA > 20 mg/ml) were Convergent: ten (29%) and nine (56%) cases respectively, [P value =0.0572]. The majority of mutations carriers (68%) have T1 and T2 pathological tumor stages, eleven cases at T1 and 12 cases in T2. Twenty-six out of 34 (76%) patients were diagnosed with prostate cancer at the age of 60 years old or more, while only five were diagnosed at age of 60 years or less. Whilst twelve out of 16 (75%) of noncarriers were diagnosed at the age of 60 or more, [p value =0.8745]. Seventeen cases were treated with radical prostatectomy, twelve of them (35%) found as high mutations carriers, and five (31%) cases were noncarriers. [P value =0.7175].

Protein sequence and mutational positions were submitted to SIFT Blink server, the server predicts if the amino acid substitution alters the protein function. The intolerant range ≤ 0.05 has been used as a limit of variants classification. More than this limit indicates as damaging to the protein function. 2 (50%) out of 5 missense variants were predicted to affect the protein function namely: (Arg70Pro) and (Glu112Lys). three of the missense variants tested (60%) were predicted as tolerated (Asn72Thr, Phe125Leu and Asp130Asn). Another important function and structure of a human protein prediction tool is PolyPhen2 HumDiv. Every time, the full protein sequence was provided with one certain amino acid substitution to be investigated. PolyPhen scores comprise a range from zero to one. 1.00 Score means that substitution is probably damaging, low scores considered a benign effect. Out of 5 missense variants, 3 (60%) were predicted as probably damaging with values range: (0.998-1.00) namely: (Arg70Pro), (Asn72Thr), and (Phe125Leu), one missense variant (Glu112Lys) possibly damaging the protein with value: (0.835), one variant (Asp130Asn) were benign (0.311). The results of missense variants effect prediction are given in Table 4.

**Table3:** Tumor pathological characteristics of *SPOP* common mutations carriers and noncarriers patients of prostate cancer.

| Pathological parameters | Carriers *<br>(n=34) | Non carriers<br>(n=16) | P** |
| --- | --- | --- | --- |
| **Pathological Gleason score** |  |  | 0.1944 |
| <7 | 9 | 3 |  |
| 7 (3 + 4) | 5 | 2 |  |
| 7 (4 + 3) | 6 | 1 |  |
| >7 | 3 | 5 |  |
| Unknown | 11 | 5 |  |
| **Prostate specific antigen** |  |  | 0.0572 |
| PSA < 10 | 9 | 1 |  |
| PSA 10-20 | 10 | 2 |  |
| PSA > 20 | 10 | 9 |  |
| Unknown | 5 | 3 |  |
| **Pathological T stage** |  |  | 0.2157 |
| pT1 | 11 | 3 |  |
| pT2 x | 12 | 4 |  |
| pT3 x | 2 | 1 |  |
| pT4 | 2 | 4 |  |
| Unknown | 7 | 4 |  |
| **Age at diagnosis/surgery** |  |  | 0.8745 |
| ≤60 years | 5 | 2 |  |
| >60 years | 26 | 12 |  |
| Unknown | 3 | 4 |  |
| **Radical prostatectomy** |  |  | 0.7175 |
| Yes | 12 | 5 |  |
| No | 13 | 7 |  |
| Unkown | 9 | 4 |  |

\*Based on carriers of one common mutation or more. \*\*Chi-squared test or Fisher's exact test (excludes missing data).

**Table 4:** Predictions results of novel missense variants effect on SPOP protein by SIFT and PolyPhen2. The variants predicted as deleterious by SIFT or PolyPhen2 in bold. The variants located in MATH domain and at the same time predicted as deleterious by the two tools; were underlined.

| CDS | Protein change | SIFT |  | PolyPhen2 |  |
| --- | --- | --- | --- | --- | --- |
| c.209G>C | <u>Arg70Pro</u> | **Affect protein** | 0.03 | **Probably damaging** | 1.000 |
| c.215A>C | Asn72Thr | Tolerated | 0.29 | **Probably damaging** | 0.998 |
| c.334G>A | <u>Glu112Lys</u> | **Affect protein** | 0.05 | **Possibly damaging** | 0.835 |
| c.373T>C | Phe125Leu | Tolerated | 0.64 | **Probably damaging** | 1.000 |
| c.388G>A | Asp130Asn | Tolerated | 0.46 | Benign | 0.311 |

## Discussion and conclusions

The finding revealed novel alterations in the conserved regions of the *SPOP* gene, one of the most mutant genes in prostate cancer through different populations. Among 50 biopsies, 34 samples have one or of high frequent mutations. This is the first time to study *SPOP* alterations in this population. The correlations between *SPOP* high mutant tissues and mutations noncarriers of prostate cancer patients and tumor characteristics were not significantly different. No statistical indications that the stage of tumor or Gleason score or PSA concentration might correlate with the presence of high mutations in the prostate tissue. This conjugate with relations between SPOP alterations and specific types of hereditary prostate cancer previously described [16]. Mutations of *SPOP* in prostate cancer recently revealed missense mutations in the MATH domain, some missense mutations were frequent fifty percent, in all cases, the *SPOP* found highly mutated in prostate cancer patients [2; 17]. This study confirmed previous findings and highlighted new mutations along with all MATH domain sequences of the *SPOP* gene. The mutations located in the conserved domain MATH are shown in Figure 1. Two subsequent serious frameshift mutations (255delA and 247delT) with high-frequency rates were detected. These two frameshift mutations probably affect MATH domain functions of interaction with Cullin3-based E3-ubiquitin ligase (Cul3), which is be found to be expressed in prostate tissues, Besides impairing other vital mechanisms of *SPOP* protein [18–21].

**Figure1:**
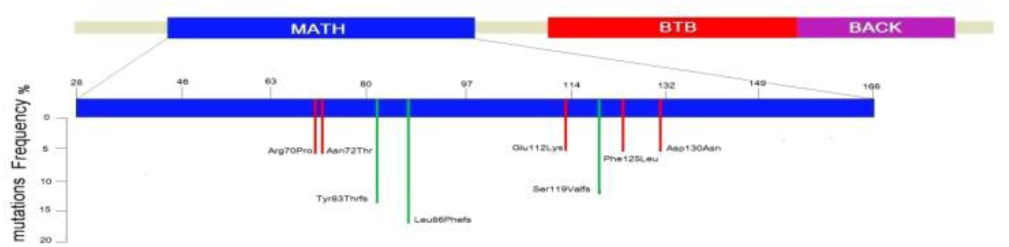
High frequent variants detected in conserved MATH domain of *SPOP* in prostate cancer biopsies. Red (short) lines indicate missense mutations; green (long) lines indicate frameshift mutations. The scheme was drawn based on: NCBI-GenBank ID: CAA04199, *SPOP* protein [*Homo sapiens*], and UniProt ID: UniProtKB - O43791 (SPOP_HUMAN).

Fifty percent of missense variants detected in this study were predicted to alter the function of vital functions of the *SPOP* protein. Prediction of missense variant by two of most powerful predictions tool: *SIFT* and *PolyPhen*2; revealed the effects of every variants on the *SPOP* protein which probably affect the function of *SPOP* leading to a defects in protein pathway as tumor suppressor genes. Notably, two missense variants located in *MATH* domain were predicted by SIFT and PolyPhen2 together to affect the protein, namely: Arg70Pro and Glu112Lys. The mutations mentioned above possibly alter *SPOP* protein vital interactions which might lead to cancer development. And less frequent missense (6%) effectively located in the MATH domain but the significant effects of these missense mutations on the biochemical and physical structure and vital functions of *SPOP* protein are uncertain. these novel mutations are not excluded to the specific population, but a confirmation of our findings in other populations will be interesting. Future examinations of such studies should concern the family history of the patients; the matter was not possible for our analysis. Biochemical and structural characteristics of *SPOP* protein after mutations highlighted in this study; also need to be investigated.

## Data Availability

All data generated or analysed during this study are included in this published article (and its supplementary information files).The datasets generated during and/or analysed during the current study are available from the corresponding author

## Acknowledgments

We are grateful to the Biopsies donors and their families, who contributed to this study, members of Virology, Oncology and Medical Biotechnology team at FSTM-Hassan II University of Casablanca, for their support us during practical stages on this study.

## Ethics approvals

Required Ethical standards were considered according to the committee of biomedical research ethics in Morocco approvals (No.: 3/2018/30 April/2018).

## Authors Contribution

Conceiving and design of the experiments: B.A, A.L,MN. B. and MM.E.Optimization the experimental approach: B.A, M.Mz and MM.E. Experiments performance: B.A. Sequencing and data analyses: B.A.,S.B.K. Sample collections, management, Technical support and proof reading:: M.Mr, A.L., A.A. Technical support and proof reading: M.Mr, A.L., A.A. Manuscript writing: B.A., MM.E. All authors approved the final version.

## References

1. Ferlay, J., Colombet, M., Soerjomataram, I., Mathers, C., Parkin, D. M., Piñeros, M., … & Bray, F.. Estimating the global cancer incidence and mortality in 2018: GLOBOCAN sources and methods. International journal of cancer 2019; 144(8), 1941–1953.

2. Blattner, M., Lee, D. J., O’Reilly, C., Park, K., MacDonald, T. Y., Khani, F., … & Heguy, A. SPOP mutations in prostate cancer across demographically diverse patient cohorts. Neoplasia (New York, NY) 2014; 16(1), 14.

3. Mani, R. S. The emerging role of speckle-type POZ protein (SPOP) in cancer development. Drug discovery today 2014; 19(9), 1498–1502,.

4. García-Flores, M., Casanova-Salas, I., Rubio-Briones, J., Calatrava, A., Domínguez-Escrig, J., Rubio, L., … & López-Guerrero, J. A. Clinico-pathological significance of the molecular alterations of the SPOP gene in prostate cancer. European journal of cancer 2014; 50(17), 2994–3002,.

5. Theurillat, J. P. P., Udeshi, N. D., Errington, W. J., Svinkina, T., Baca, S. C., Pop, M., … & Moch, H. 2014. Ubiquitylome analysis identifies dysregulation of effector substrates in SPOP-mutant prostate cancer. Science 2014; 346(6205), 85–89,.

6. Li, C., Ao, J., Fu, J., Lee, D. F., Xu, J., Lonard, D., & O’Malley, B. W. Tumor-suppressor role for the SPOP ubiquitin ligase in signal-dependent proteolysis of the oncogenic co-activator SRC-3/AIB1. Oncogene 2011; 30(42), 4350,.

7. Barbieri, C. E., Baca, S. C., Lawrence, M. S., Demichelis, F., Blattner, M., Theurillat, J. P., … & Nickerson, E. Exome sequencing identifies recurrent SPOP, FOXA1 and MED12 mutations in prostate cancer. Nature genetics, 2012; 44(6), 685,.

8. Boysen, G., Barbieri, C. E., Prandi, D., Blattner, M., Chae, S. S., Dahija, A., … & Huang, J., SPOP mutation leads to genomic instability in prostate cancer. Elife 2015; 4, e09207,

9. Gan, W., Dai, X., Lunardi, A., Li, Z., Inuzuka, H., Liu, P., … & Asara, J. M., SPOP promotes ubiquitination and degradation of the ERG oncoprotein to suppress prostate cancer progression. Molecular cell 2015; 59(6), 917–930.

10. An, J., Wang, C., Deng, Y., Yu, L., & Huang, H. Destruction of full-length androgen receptor by wild-type SPOP, but not prostate-cancer-associated mutants. Cell reports 2014; 6(4), 657–669.

11. Culp, M. B., I. Soerjomataram, J. A. Efstathiou, F. Bray, and A. Jemal. Recent Global Patterns in Prostate Cancer Incidence and Mortality Rates. European urology 2019.

12. Hijikata, R Raju, S Keerthikumar, S Ramabadran, L Balakrishnan, SK Ramadoss, A Pandey, S Mohan, O Ohara., Mutation at Glance: an integrative web application for analysing mutations from human genetic diseases. DNA Res 2010; 17, 3, 197–208,.

13. Wildeman M, van Ophuizen E, den Dunnen JT, Taschner PE. Improving sequence variant descriptions in mutation databases and literature using the MUTALYZER sequence variation nomenclature checker. Hum Mutat 2008; 29:6–13.

14. Kumar P, Henikoff S, Ng PC., Predicting the effects of coding non-synonymous variants on protein function using the SIFT algorithm, Nat Protoc. 2009; 4(7) 1073–1081. doi: 10.1038/nprot.2009.86 PMID:19561590.

15. Adzhubei IA, Schmidt S, Peshkin L, Ramensky VE, Gerasimova A, Bork P, et al.,. A method and server for predicting damaging missense mutations. Nat Methods 2010; 7(4) 248–249. doi: 10.1038/nmeth0410-248 PMID: 20354512.

16. Zuhlke, K. A., Johnson, A. M., Tomlins, S. A., Palanisamy, N., Carpten, J. D., Lange, E. M., … & Cooney, K. A., Identification of a novel germline SPOP mutation in a family with hereditary prostate cancer. The Prostate 2014; 74(9), 983–990.

17. Yoon, N., Sung, J. Y., Kang, S. Y., Kwon, G. Y., & Choi, Y. L., SPOP mutation in prostate cancers in Korean population: variation in its mutation frequency among ethnic groups. International journal of clinical and experimental pathology 2016; 9(3), 4123–4128.

18. Geng, C., Rajapakshe, K., Shah, S. S., Shou, J., Eedunuri, V. K., Foley, C., … & Bond, R.. Androgen receptor is the key transcriptional mediator of the tumor suppressor SPOP in prostate cancer. Cancer research, 2014; 74(19), 5631–5643

19. Bouchard, J. J., Otero, J. H., Scott, D. C., Szulc, E., Martin, E. W., Sabri, N., … & Schulman, B. A., Cancer mutations of the tumor suppressor SPOP disrupt the formation of active, phase-separated compartments. Molecular cell; 2018, 72(1) 19–36.

20. Yan, Yuqian, et al., The novel BET-CBP/p300 dual inhibitor NEO2734 is active in SPOP mutant and wild-type prostate cancer. EMBO Molecular Medicine; 2019.

21. Cuneo, Matthew J. et Mittag, Tanja., The ubiquitin ligase adaptor SPOP in cancer. The FEBS journal 2019; vol. 286, no 20 p. 3946–3958.

